## Supplementary Materials for "Machine learning approach to dynamic risk modeling of mortality in COVID-19: a UK Biobank study" (from another version of this preprint: version 1, 2021-02-10)

### Supplementary Material

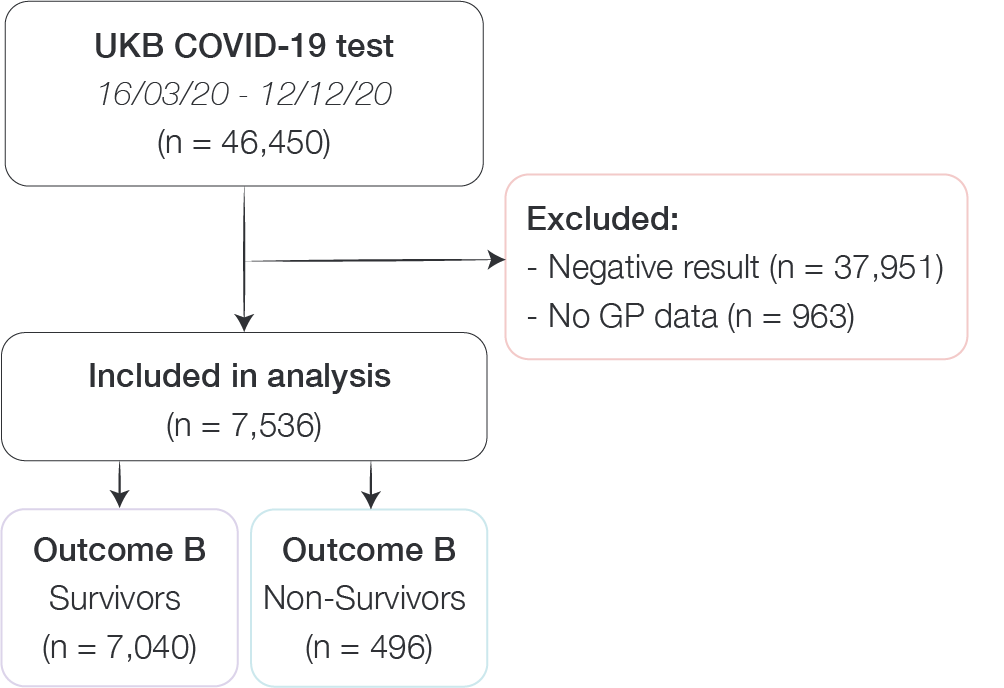

###### **Supplementary Figure 1.** Flow chart for participant selection.

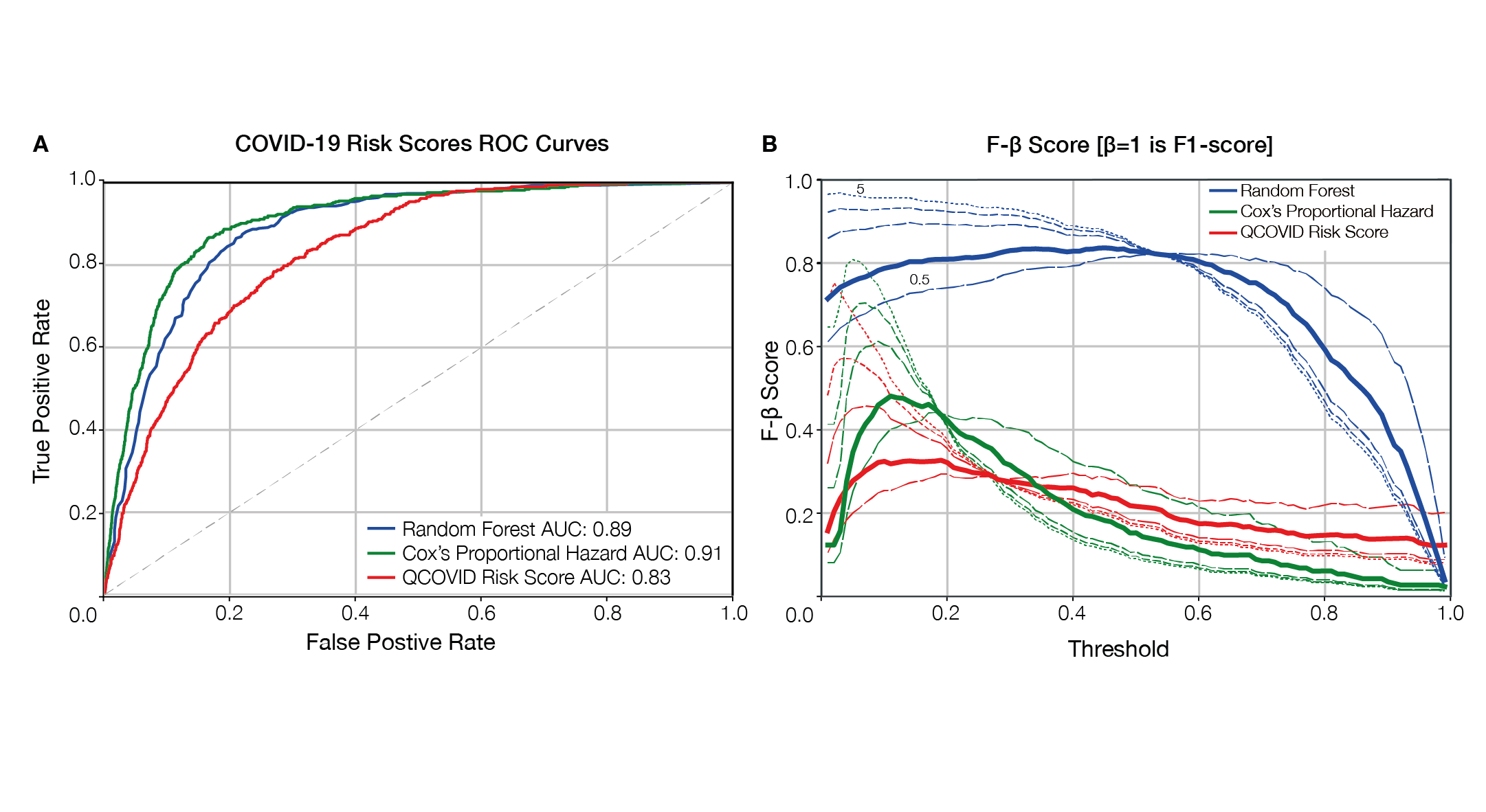

###### **Supplementary Figure 2.** Model performance evaluation without age variable included in model: (A) the receiver operating characteristic (ROC) curve comparison shown for our Random Forest (RF) and Cox models against QCOVID; (B) the F-β score generated at β=1 (F1-score in bold), β=[ 0.5, 2, 3, 5], shown in decreasing size dashed line.

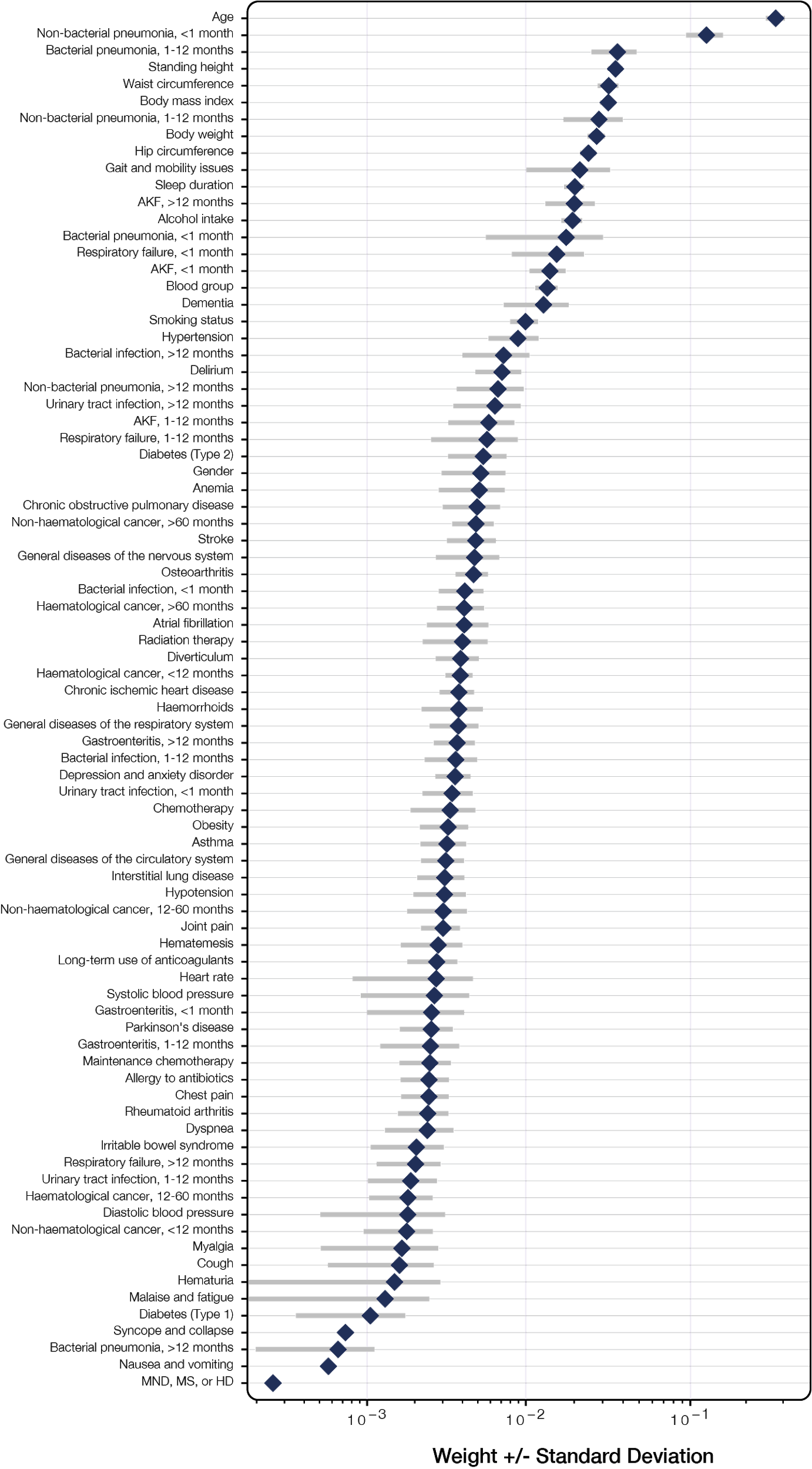

###### **Supplementary Figure 3.** Random Forest feature ranking of characteristics included in the model. AKF = acute kidney failure, MND = motor neurone disease, MS = multiple sclerosis, HD = Huntington’s Disease.

| **Feature Type** | **Feature Name** | **Generalised Coding** |
| --- | --- | --- |
| Conditions | Acute kidney failure | N17* |
| Conditions | Non-bacterial pneumonia | J12*; J17*; J181; J189 |
| Conditions | Urinary tract infection | N30*; N39.0; A56.0; A56.1; A56.2; A54.0; A54.1; N29.1 |
| Conditions | Respiratory failure | J96* |
| Conditions | Bacterial infection | A39*; A4*; A04*; A05*; Z22.3* |
| Conditions | Bacterial pneumonia | J12; J13; J15* |
| Conditions | Osteoarthritis | M17*; M18*; M19*; M15*; M16* |
| Conditions | Hypertension | I10* |
| Conditions | Depression and anxiety disorder | F32*; F33*; F41*; F43* |
| Conditions | Dementia | F00*; F01*; F02*; F03*; G309 |
| Conditions | Gastroenteritis | A09*; K29.7; K52* |
| Conditions | Diverticulum | K57*; Q43.0; K31.4; N32.3; |
| Conditions | Diabetes (Type 2) | E11* |
| Conditions | Anemia | D50*; D51*; D52*; D53*; D64* |
| Conditions | Asthma | J45* J46* |
| Conditions | Haemorrhoids | K64*; I849; I842; I846 |
| Conditions | Chronic ischemic heart disease | I25* |
| Conditions | General diseases of the circulatory system | Z86.7 |
| Conditions | Atrial fibrillation | I48* |
| Conditions | General diseases of the respiratory system | Z87.0 |
| Conditions | General diseases of the nervous system | Z86.6 |
| Conditions | Haematological cancer | C81.0 - 96.9; D45-47.9 |
| Conditions | Non-haematological cancer | D37.5; C19*; C20*; C21*; D12*; C44*; C39*; C78.0; C78.1; C78.2; C78.3; C78.0; Z85.2; Z85.3 |
| Conditions | Rheumatoid arthritis | M05*; M06*; M32* |
| Conditions | Diabetes (Type 1) | E10* |
| Conditions | Stroke | I6* |
| Conditions | Chronic obstructive pulmonary disease | f42016;J44* |
| Conditions | Interstitial lung disease | J84* |
| Conditions | Irritable bowel syndrome | K58* |
| Conditions | Parkinson's disease | G20; G21; f42030 |
| Conditions | MND, MS, or HD | G10-G14; f42028 |
| Conditions | Hypotension | I95* |
| Demographic | Age | f34 |
| Demographic | Waist circumference | f48 |
| Demographic | Hip circumference | f49 |
| Demographic | Gender | f31 |
| Lifestyle / Anthropometrics | Body mass index | f21001; 60621009; 22K* |
| Lifestyle / Anthropometrics | Body weight | 27113001; 22A* |
| Lifestyle / Anthropometrics | Sleep duration | f1160 |
| Lifestyle / Anthropometrics | Alcohol intake | f1558; Z721; 136* |
| Lifestyle / Anthropometrics | Smoking status | f20116; Z720 |
| Lifestyle / Anthropometrics | Obesity | E66* |
| Lifestyle / Anthropometrics | Standing height | 248333004; f50 |
| Lifestyle / Anthropometrics | Blood group | fblood_group |
| Lifestyle / Anthropometrics | Gait and mobility issues | R26*; R27*; R29.6; R54; M62.5; W182 |
| Medication | Maintenance chemotherapy | Z51.2 |
| Medication | Allergy to antibiotics | Z88.0; Z88.1 |
| Medication | Long-term use of anticoagulants | Z92.1 |
| Medication | Radiation therapy | Z92.3 |
| Medication | Chemotherapy | Z926 |
| Symptoms | Joint pain | M25.5*; M79.6*; M79.1; M54* |
| Symptoms | Delirium | F05* |
| Symptoms | Hematemesis | K92* |
| Symptoms | Syncope and collapse | R55 |
| Symptoms | Dyspnea | R06* |
| Symptoms | Cough | R05 |
| Symptoms | Myalgia | M79.1; M79.6; M79.7; |
| Symptoms | Nausea and vomiting | R11 |
| Symptoms | Chest pain | R07* |
| Symptoms | Malaise and fatigue | R53; R40.0 |
| Symptoms | Hematuria | R31; N02* |
| Vital signs | Diastolic blood pressure | 1091811000000102; 246A* |
| Vital signs | Systolic blood pressure | 72313002; 2469 |
| Vital signs | Heart rate | 78564009; X773s; 242.. |
| Vital signs | Body temperature | 415974002; X75Xk |
| Vital signs | Oxygen saturation | X770D; X7708 |
| Vital signs | Respiratory rate | X774f; 235* |

###### **Supplementary Table 1.** Codings for characteristics reported in the model. Each source has unique coding attributes of patient characteristics, pre-existing clinical conditions, symptoms, and vital signs. For example, Primary Care (GP) dataset includes coding classifications of SNOMED CT, local EMIS, TPP and CTV3 codes depending on the source of the data point. * = all fields that contain string; f = UK Biobank field ID. MND = motor neurone disease, MS = multiple sclerosis, HD = Huntington’s Disease.

| **QRisk Variable** | **Biobank FieldID** | **ICD10 Code** |
| --- | --- | --- |
| Sex | 31 | - |
| Age | 21022 | - |
| Townsend material deprivation score | 189 | - |
| White | 21000 | - |
| Indian | 21000 | - |
| Pakistani | 21000 | - |
| Bangladeshi | 21000 | - |
| Other Asian | 21000 | - |
| Caribbean | 21000 | - |
| Black African | 21000 | - |
| Chinese | 21000 | - |
| Other ethnic group | 21000 | - |
| BMI | 21001; 21002 | - |
| Smoking status | 20116 | - |
| Not in care home or homeless (defined as missing) | - | - |
| Lives in residential or nursing home | - | Y92.12 |
| Lives in residential or nursing home | - | Z59.0 |
| No learning disability (defined as missing) | - | - |
| Learning disability apart from Down's syndrome |  | F7*; F81.9 |
| Down's syndrome | 41270 | Q90* |
| No Kidney Failure | 41270 | - |
| Chronic kidney disease stage 3 | 41270 | N18.3 |
| Chronic kidney disease stage 4 | 41270 | N18.4 |
| Chronic kidney disease stage 5 | 41270 | N18.5 |
| Chronic kidney disease stage 5 with dialysis | 41270 | N18.6 |
| Chronic kidney disease stage 5 with transplant | 41270 | N18.5 |
| Not on chemotherapy in past 12 months | - | Z51.1 |
| Blood cancer |  | C81.0 - 96.9; D45-47.9 |
| Bone marrow or stem cell transplant in past 6 months | 41270 | Z94.8 |
| Respiratory tract cancer | 41270 | C39*; C78.0; C78.1; C78.2; C78.3; C78.0; Z85.2 |
| Radiotherapy in past 6 months | 41270 | Z51.0 |
| Solid organ transplant (excluding kidney and bone marrow) | 41270 | Z94* (NOT Z94.0 or Z94.81) |
| Sickle cell disease or severe immunodeficiency | 41270 | D57 |
| No diabetes | 41270 | E10-E14 |
| Type 1 diabetes | 41270 | E10* |
| Type 2 diabetes | 41270 | E11* |
| Chronic obstructive pulmonary disease | 42016 | J44* |
| Asthma | 42014 | J45* |
| Rare lung conditions (bronchiectasis, cystic fibrosis, or alveolitis) | 41270 | J47*; E84*; J84*; J82* |
| Pulmonary hypertension or pulmonary fibrosis | 41270 | I27*; J84.1 |
| Coronary heart disease |  | I20-I25 |
| Stroke | 42006 | I6* |
| Atrial fibrillation | 131350; 20002 | I48* |
| Congestive cardiac failure | 41270; 131354 | I50* |
| Thrombo-embolism | 41270 | I80.0-I80.3; I80.8-I80.9; I82.9; O22.2 – O22.3; O87.0 –  O87.1; I26.0; I26.9 |
| Peripheral vascular disease | 41270 | I73* |
| Congenital heart disease | 41270 | Q24* |
| Dementia | 42018 | F00-F09 |
| Parkinson's disease | 42030 | G20; G21 |
| Epilepsy | 41270 | G40* |
| Motor neurone disease, multiple sclerosis, myaesthenia gravis, or Huntington's Disease | 42028 | G10-G14 |
| Cerebral palsy | 41270 | G80* |
| Severe mental illness |  | F2*; F3*; F4*; F5*; F6* |
| Osteoporotic fracture (hip, spine, wrist, humerus) | 41270 | M80* |
| Rheumatoid arthritis or SLE | 41270 | M32*;M06*; M05* |
| Cirrhosis of liver | 41270 | K74* |
| Not included |  |  |
| Chemotherapy grade A | - | - |
| Chemotherapy grade B | - | - |
| Chemotherapy grade C | - | - |
| Immunosuppressant medication from GP 4+ scripts in past 6 months | - | - |
| Leukotriene or long acting β-agonist 4+ scripts in past 6 months | - | - |
| Oral steroids 4+ scripts in past 6 months | - | - |

###### **Supplementary Table 2.** Coding mapping for QCOVID to UK Biobank dataset and ICD-10 codes for available hospitalisation data. * = all fields that contain string.

| **Feature name** | **Category** | **coef** | **exp(coef)** | **se(coef)** | **coef lower 95%** | **coef upper 95%** | **exp(coef) lower 95%** | **exp(coef) upper 95%** | **z** | **p** | **-log2(p)** |
| --- | --- | --- | --- | --- | --- | --- | --- | --- | --- | --- | --- |
| Maintenance chemotherapy | Medication | 0.04 | 1.05 | 0.09 | -0.13 | 0.22 | 0.88 | 1.24 | 0.51 | 0.61 | 0.71 |
| Age | Demographic | 1.28 | 3.58 | 0.22 | 0.84 | 1.71 | 2.32 | 5.54 | 5.74 | <0.005 | 26.65 |
| Body mass index | Lifestyle / Anthropometrics | 0.16 | 1.17 | 0.24 | -0.32 | 0.63 | 0.72 | 1.89 | 0.64 | 0.53 | 0.93 |
| Waist circumference | Demographic | 0.49 | 1.64 | 0.25 | 0.01 | 0.98 | 1.01 | 2.66 | 1.98 | 0.05 | 4.39 |
| Hip circumference | Demographic | 0.03 | 1.03 | 0.33 | -0.62 | 0.68 | 0.54 | 1.97 | 0.08 | 0.93 | 0.1 |
| Body weight | Lifestyle / Anthropometrics | 0.11 | 1.12 | 0.2 | -0.27 | 0.5 | 0.76 | 1.64 | 0.58 | 0.56 | 0.83 |
| Sleep duration | Lifestyle / Anthropometrics | 0.22 | 1.24 | 0.29 | -0.35 | 0.78 | 0.71 | 2.18 | 0.75 | 0.45 | 1.14 |
| Alcohol intake | Lifestyle / Anthropometrics | 0 | 1 | 0 | 0 | 0 | 1 | 1 | 0 | 1 | 0 |
| Smoking status | Lifestyle / Anthropometrics | -0.06 | 0.94 | 0.08 | -0.22 | 0.11 | 0.8 | 1.11 | -0.68 | 0.5 | 1.01 |
| Gender | Demographic | 0.08 | 1.09 | 0.04 | 0.01 | 0.16 | 1.01 | 1.17 | 2.16 | 0.03 | 5.04 |
| Obesity | Lifestyle / Anthropometrics | 0.03 | 1.03 | 0.06 | -0.08 | 0.14 | 0.92 | 1.15 | 0.52 | 0.6 | 0.73 |
| Standing height | Lifestyle / Anthropometrics | 0 | 1 | 0 | 0 | 0 | 1 | 1 | 0 | 1 | 0 |
| Osteoarthritis | Conditions | 0.01 | 1.01 | 0.04 | -0.08 | 0.1 | 0.93 | 1.1 | 0.25 | 0.8 | 0.32 |
| Hypertension | Conditions | 0.12 | 1.12 | 0.04 | 0.04 | 0.19 | 1.04 | 1.22 | 2.89 | <0.005 | 8.02 |
| Gait and mobility issues | Lifestyle / Anthropometrics | 0.23 | 1.26 | 0.06 | 0.11 | 0.35 | 1.11 | 1.42 | 3.69 | <0.005 | 12.14 |
| Depression and anxiety disorder | Conditions | 0.01 | 1.01 | 0.06 | -0.09 | 0.12 | 0.91 | 1.13 | 0.24 | 0.81 | 0.31 |
| Allergy to antibiotics | Medication | 0 | 1 | 0.06 | -0.12 | 0.12 | 0.89 | 1.13 | -0.01 | 0.99 | 0.01 |
| Dementia | Conditions | 0.29 | 1.34 | 0.08 | 0.13 | 0.45 | 1.14 | 1.56 | 3.62 | <0.005 | 11.75 |
| Diverticulum | Conditions | 0 | 1 | 0 | 0 | 0 | 1 | 1 | 0 | 1 | 0 |
| Diabetes (Type 2) | Conditions | 0.13 | 1.14 | 0.06 | 0.02 | 0.24 | 1.02 | 1.27 | 2.33 | 0.02 | 5.67 |
| Anemia | Conditions | 0.13 | 1.14 | 0.06 | 0.02 | 0.24 | 1.02 | 1.28 | 2.33 | 0.02 | 5.66 |
| Asthma | Conditions | -0.01 | 0.99 | 0.05 | -0.12 | 0.09 | 0.89 | 1.1 | -0.25 | 0.8 | 0.31 |
| Long-term use of anticoagulants | Medication | 0.08 | 1.09 | 0.06 | -0.04 | 0.21 | 0.96 | 1.23 | 1.38 | 0.17 | 2.56 |
| Haemorrhoids | Conditions | -0.05 | 0.95 | 0.06 | -0.18 | 0.07 | 0.84 | 1.08 | -0.81 | 0.42 | 1.27 |
| Chronic ischemic heart disease | Conditions | 0.08 | 1.08 | 0.05 | -0.03 | 0.18 | 0.97 | 1.2 | 1.41 | 0.16 | 2.65 |
| General diseases of the circulatory system | Conditions | 0.06 | 1.07 | 0.06 | -0.05 | 0.18 | 0.95 | 1.19 | 1.09 | 0.28 | 1.86 |
| Atrial fibrillation | Conditions | 0.11 | 1.11 | 0.06 | -0.01 | 0.23 | 0.99 | 1.25 | 1.73 | 0.08 | 3.59 |
| General diseases of the respiratory system | Conditions | 0 | 1 | 0 | 0 | 0 | 1 | 1 | 0 | 1 | 0 |
| Radiation therapy | Medication | 0.05 | 1.05 | 0.12 | -0.17 | 0.28 | 0.84 | 1.32 | 0.45 | 0.65 | 0.62 |
| General diseases of the nervous system | Conditions | 0.06 | 1.06 | 0.08 | -0.09 | 0.21 | 0.92 | 1.24 | 0.82 | 0.42 | 1.27 |
| Chemotherapy | Medication | 0.08 | 1.09 | 0.12 | -0.15 | 0.31 | 0.86 | 1.36 | 0.71 | 0.48 | 1.07 |
| Hypotension | Conditions | 0.13 | 1.14 | 0.09 | -0.05 | 0.32 | 0.95 | 1.37 | 1.4 | 0.16 | 2.63 |
| Joint pain | Symptoms | 0.03 | 1.03 | 0.06 | -0.09 | 0.14 | 0.91 | 1.15 | 0.45 | 0.66 | 0.61 |
| Delirium | Symptoms | 0.22 | 1.24 | 0.1 | 0.02 | 0.42 | 1.02 | 1.52 | 2.11 | 0.03 | 4.86 |
| Hematemesis | Symptoms | 0 | 1 | 0 | 0 | 0 | 1 | 1 | 0 | 1 | 0 |
| Syncope and collapse | Symptoms | 0 | 1 | 0 | -0.01 | 0.01 | 0.99 | 1.01 | 0 | 1 | 0 |
| Dyspnea | Symptoms | 0.09 | 1.09 | 0.11 | -0.13 | 0.32 | 0.87 | 1.37 | 0.79 | 0.43 | 1.22 |
| Cough | Symptoms | 0.06 | 1.06 | 0.2 | -0.33 | 0.44 | 0.72 | 1.56 | 0.29 | 0.77 | 0.38 |
| Myalgia | Symptoms | -0.02 | 0.98 | 0.12 | -0.26 | 0.22 | 0.77 | 1.24 | -0.19 | 0.85 | 0.24 |
| Nausea and vomiting | Symptoms | 0.26 | 1.29 | 0.26 | -0.25 | 0.76 | 0.78 | 2.13 | 1 | 0.32 | 1.65 |
| Chest pain | Symptoms | 0 | 1 | 0 | 0 | 0 | 1 | 1 | 0 | 1 | 0 |
| Diastolic blood pressure | Vital signs | 0 | 1 | 0 | 0 | 0 | 1 | 1 | 0 | 1 | 0 |
| Systolic blood pressure | Vital signs | 0 | 1 | 0 | 0 | 0 | 1 | 1 | 0 | 1 | 0 |
| Heart rate | Vital signs | 0 | 1 | 0 | 0 | 0 | 1 | 1 | -0.06 | 0.95 | 0.08 |
| Malaise and fatigue | Symptoms | 0 | 1 | 0 | 0 | 0 | 1 | 1 | 0 | 1 | 0 |
| Rheumatoid arthritis | Conditions | 0.13 | 1.14 | 0.11 | -0.08 | 0.34 | 0.92 | 1.4 | 1.21 | 0.22 | 2.15 |
| Diabetes (Type 1) | Conditions | 0.21 | 1.23 | 0.15 | -0.07 | 0.5 | 0.93 | 1.64 | 1.44 | 0.15 | 2.75 |
| Stroke | Conditions | 0.11 | 1.12 | 0.07 | -0.02 | 0.24 | 0.98 | 1.27 | 1.6 | 0.11 | 3.18 |
| Chronic obstructive pulmonary disease | Conditions | 0.14 | 1.15 | 0.07 | 0 | 0.28 | 1 | 1.33 | 2.01 | 0.04 | 4.49 |
| Interstitial lung disease | Conditions | 0.33 | 1.39 | 0.16 | 0.02 | 0.64 | 1.02 | 1.9 | 2.1 | 0.04 | 4.8 |
| Irritable bowel syndrome | Conditions | 0 | 1 | 0.1 | -0.19 | 0.19 | 0.83 | 1.21 | 0.01 | 0.99 | 0.02 |
| Parkinson's disease | Conditions | 0.29 | 1.34 | 0.14 | 0.02 | 0.56 | 1.02 | 1.75 | 2.11 | 0.03 | 4.85 |
| MND, MS, or Huntington's | Conditions | 0 | 1 | 0.01 | -0.01 | 0.01 | 0.99 | 1.01 | 0 | 1 | 0 |
| Hematuria | Symptoms | 0 | 1 | 0.26 | -0.51 | 0.52 | 0.6 | 1.68 | 0.02 | 0.99 | 0.02 |
| Blood group | Lifestyle / Anthropometrics | 0 | 1 | 0.06 | -0.12 | 0.12 | 0.89 | 1.13 | 0.01 | 0.99 | 0.01 |
| Haematological cancer, <12 months | Conditions | 0.44 | 1.56 | 0.16 | 0.12 | 0.76 | 1.13 | 2.15 | 2.69 | 0.01 | 7.14 |
| Haematological cancer, 12-60 months | Conditions | 0.23 | 1.25 | 0.18 | -0.12 | 0.57 | 0.89 | 1.78 | 1.28 | 0.2 | 2.32 |
| Haematological cancer, >60 months | Conditions | 0.27 | 1.31 | 0.17 | -0.06 | 0.6 | 0.94 | 1.82 | 1.59 | 0.11 | 3.15 |
| Non-haematological cancer, <12 months | Conditions | 0.15 | 1.16 | 0.13 | -0.11 | 0.42 | 0.89 | 1.51 | 1.13 | 0.26 | 1.95 |
| Non-haematological cancer, 12-60 months | Conditions | -0.02 | 0.98 | 0.08 | -0.18 | 0.15 | 0.84 | 1.16 | -0.19 | 0.85 | 0.24 |
| Non-haematological cancer, >60 months | Conditions | 0 | 1 | 0 | 0 | 0 | 1 | 1 | 0 | 1 | 0 |
| Urinary tract infection, <1 month | Conditions | 0.11 | 1.12 | 0.16 | -0.21 | 0.43 | 0.81 | 1.54 | 0.7 | 0.49 | 1.04 |
| Urinary tract infection, 1-12 months | Conditions | 0.11 | 1.12 | 0.13 | -0.15 | 0.37 | 0.86 | 1.45 | 0.85 | 0.4 | 1.34 |
| Urinary tract infection, >12 months | Conditions | 0.16 | 1.17 | 0.07 | 0.03 | 0.29 | 1.03 | 1.34 | 2.42 | 0.02 | 6 |
| Respiratory failure, <1 month | Conditions | 0.5 | 1.64 | 0.09 | 0.32 | 0.68 | 1.37 | 1.97 | 5.38 | <0.005 | 23.69 |
| Respiratory failure, 1-12 months | Conditions | 0.26 | 1.3 | 0.12 | 0.02 | 0.5 | 1.02 | 1.65 | 2.12 | 0.03 | 4.89 |
| Respiratory failure, >12 months | Conditions | 0.14 | 1.15 | 0.13 | -0.12 | 0.41 | 0.89 | 1.5 | 1.07 | 0.29 | 1.81 |
| AKF, <1 month | Conditions | 0.53 | 1.7 | 0.1 | 0.34 | 0.72 | 1.4 | 2.06 | 5.43 | <0.005 | 24.06 |
| AKF, 1-12 months | Conditions | 0.34 | 1.41 | 0.1 | 0.14 | 0.54 | 1.15 | 1.72 | 3.34 | <0.005 | 10.22 |
| AKF, >12 months | Conditions | 0.26 | 1.3 | 0.09 | 0.09 | 0.43 | 1.09 | 1.54 | 3.01 | <0.005 | 8.58 |
| Non-bacterial pneumonia, <1 month | Conditions | 0.46 | 1.58 | 0.06 | 0.33 | 0.58 | 1.39 | 1.79 | 7.14 | <0.005 | 39.99 |
| Non-bacterial pneumonia, 1-12 months | Conditions | 0.26 | 1.3 | 0.08 | 0.11 | 0.42 | 1.11 | 1.52 | 3.29 | <0.005 | 9.97 |
| Non-bacterial pneumonia, >12 months | Conditions | 0.1 | 1.1 | 0.07 | -0.05 | 0.24 | 0.95 | 1.28 | 1.34 | 0.18 | 2.46 |
| Bacterial pneumonia, <1 month | Conditions | 0.46 | 1.58 | 0.07 | 0.33 | 0.59 | 1.39 | 1.8 | 6.92 | <0.005 | 37.66 |
| Bacterial pneumonia, 1-12 months | Conditions | 0.32 | 1.38 | 0.09 | 0.14 | 0.5 | 1.15 | 1.65 | 3.43 | <0.005 | 10.69 |
| Bacterial pneumonia, >12 months | Conditions | 0.02 | 1.02 | 0.25 | -0.47 | 0.51 | 0.62 | 1.67 | 0.09 | 0.93 | 0.1 |
| Bacterial infection, <1 month | Conditions | 0.29 | 1.34 | 0.12 | 0.06 | 0.53 | 1.06 | 1.7 | 2.46 | 0.01 | 6.18 |
| Bacterial infection, 1-12 months | Conditions | 0.15 | 1.16 | 0.11 | -0.08 | 0.37 | 0.93 | 1.45 | 1.29 | 0.2 | 2.34 |
| Bacterial infection, >12 months | Conditions | 0.1 | 1.11 | 0.08 | -0.06 | 0.27 | 0.94 | 1.31 | 1.23 | 0.22 | 2.18 |
| Gastroenteritis, <1 month | Conditions | 0 | 1 | 0 | 0 | 0 | 1 | 1 | 0 | 1 | 0 |
| Gastroenteritis, 1-12 months | Conditions | 0.05 | 1.06 | 0.14 | -0.22 | 0.32 | 0.81 | 1.38 | 0.39 | 0.7 | 0.52 |
| Gastroenteritis, >12 months | Conditions | 0 | 1 | 0 | 0 | 0 | 1 | 1 | 0 | 1 | 0 |

###### **Supplementary Table 3.** Cox Proportional Hazards model coefficients of COVID-19 mortality in UK Biobank cohort. AKF = acute kidney failure, MND = motor neurone disease, MS = multiple sclerosis, HR = hazard ratio, CI = confidence interval.

| **Section/Topic** | **Item** |  | **Checklist Item** | **Page** |
| --- | --- | --- | --- | --- |
| Title and abstract | | | | |
| Title | 1 | D;V | Identify the study as developing and/or validating a multivariable prediction model, the target population, and the outcome to be predicted.  Machine learning approach to dynamic risk modelling of mortality in COVID-19: a UK Biobank cohort study  Identifies the target population (COVID-19 Positive UK Biobank patients), outcome (Mortality) and the prediction model (Machine Learning) | Title page |
| Abstract | 2 | D;V | Provide a summary of objectives, study design, setting, participants, sample size, predictors, outcome, statistical analysis, results, and conclusions. | Summary paragraph |
| Introduction | | | | |
| Background and objectives | 3a | D;V | Explain the medical context (including whether diagnostic or prognostic) and rationale for developing or validating the multivariable prediction model, including references to existing models. | 2 |
|  | 3b | D;V | Specify the objectives, including whether the study describes the development or validation of the model or both. | 2 |
| Methods | | | | |
| Source of data | 4a | D;V | Describe the study design or source of data (e.g., randomized trial, cohort, or registry data), separately for the development and validation data sets, if applicable.  Cohort of COVID-19+ participants in UK Biobank, linked to Electronic Health Records. | 11 |
|  | 4b | D;V | Specify the key study dates, including start of accrual; end of accrual; and, if applicable, end of follow-up.  COVID-19 test result data were available for the period 16^th^ March 2020 to 12^th^ December 2020 | 11 |
| Participants | 5a | D;V | Specify key elements of the study setting (e.g., primary care, secondary care, general population) including number and location of centres.  UK Biobank data, COVID-19 results, hospital admission, primary care, and death records | 11 |
|  | 5b | D;V | Describe eligibility criteria for participants.  Patients in UK Biobank with a positive SARS-CoV2 RT-PCR and linked primary care records. | 11 |
|  | 5c | D;V | Give details of treatments received, if relevant.  Non-applicable | - |
| Outcome | 6a | D;V | Clearly define the outcome that is predicted by the prediction model, including how and when assessed.  Mortality (death) at least 7 days after COVID-19 positive test results | 11-13 |
|  | 6b | D;V | Report any actions to blind assessment of the outcome to be predicted. | N/A |
| Predictors | 7a | D;V | Clearly define all predictors used in developing or validating the multivariable prediction model, including how and when they were measured.  Predictors included available patient demographics, lifestyle factors, medical history, vitals and symptoms in primary care records +/- two weeks of COVID-19 positive test results. These were developed based on data-driven modelling and refinement from a team of clinicians as to what was relevant and comprehensive. | Table 1 |
|  | 7b | D;V | Report any actions to blind assessment of predictors for the outcome and other predictors. | N/A |
| Sample size | 8 | D;V | Explain how the study size was arrived at.  The sample size differed based on time window and data availability within the UK Biobank | S Fig 1 |
| Missing data | 9 | D;V | Describe how missing data were handled (e.g., complete-case analysis, single imputation, multiple imputation) with details of any imputation method.  Missing values were substituted with the mean value in the UK Biobank | Fig 1 |
| Statistical analysis methods | 10a | D | Describe how predictors were handled in the analyses.  All predictors were used in the model for prediction. | 11-13 |
|  | 10b | D | Specify type of model, all model-building procedures (including any predictor selection), and method for internal validation.  Models used in this study included a random forest model and a Cox proportional hazard model. | 11-13 |
|  | 10c | V | For validation, describe how the predictions were calculated.  Leave-one-out (LOO) experiment was carried out | 12-13 |
|  | 10d | D;V | Specify all measures used to assess model performance and, if relevant, to compare multiple models.  Measures used to compare models included model accuracy, sensitivity, specificity, AUC-ROC,AUC-PRC, and F1-statistic. | 12-13 |
|  | 10e | V | Describe any model updating (e.g., recalibration) arising from the validation, if done.  No model recalibration was performed after training. The model was tested without Age to check for overfitting of the variable. | 12-13 |
| Risk groups | 11 | D;V | Provide details on how risk groups were created, if done.  No risk groups were created. | N/A |
| Development vs. validation | 12 | V | For validation, identify any differences from the development data in setting, eligibility criteria, outcome, and predictors.  For validation, there are no differences from development data in setting, eligibility, outcome, predictors or otherwise. | N/A |
| Results | | | | |
| Participants | 13a | D;V | Describe the flow of participants through the study, including the number of participants with and without the outcome and, if applicable, a summary of the follow-up time. A diagram may be helpful.  The number of patients without the outcome differed based on experiment and based on time window. | S Fig 1 |
|  | 13b | D;V | Describe the characteristics of the participants (basic demographics, clinical features, available predictors), including the number of participants with missing data for predictors and outcome.  Demographics, lifestyle factors, medical history, symptoms and vital signs. | Table 1 |
|  | 13c | V | For validation, show a comparison with the development data of the distribution of important variables (demographics, predictors and outcome).  For validation, there are no differences from development data in setting, eligibility, outcome, predictors or otherwise. | Table 1 |
| Model development | 14a | D | Specify the number of participants and outcome events in each analysis.  We show the number of patients involved and the proportion of events in each experiment by time window in Table 1. | Table 1 |
|  | 14b | D | If done, report the unadjusted association between each candidate predictor and outcome.  N/A | N/A |
| Model specification | 15a | D | Present the full prediction model to allow predictions for individuals (i.e., all regression coefficients, and model intercept or baseline survival at a given time point).  The Cox model outcomes are presented in figure 3. See supplementary figure/table for further information. | Fig 3 |
|  | 15b | D | Explain how to the use the prediction model.  The prediction model can be used in a patient-facing questionnaire and may be deployed on mobile/microsite. | S Fig 3 |
| Model performance | 16 | D;V | Report performance measures (with CIs) for the prediction model.  Please refer to Figure 2 for model performance | S Table 3 |
| Model-updating | 17 | V | If done, report the results from any model updating (i.e., model specification, model performance).  No model recalibration was performed after training. The model was tested without Age to check for overfitting of the variable. | See discussion 7-10 |
| Discussion | | | | |
| Limitations | 18 | D;V | Discuss any limitations of the study (such as non-representative sample, few events per predictor, missing data).  Details on study limitations include limited age range, missingness present in data, inherent limitations using EHRs and temporal aspects of data. | 10 |
| Interpretation | 19a | V | For validation, discuss the results with reference to performance in the development data, and any other validation data.  Please see the manuscript for full discussion. | 7-10 |
|  | 19b | D;V | Give an overall interpretation of the results, considering objectives, limitations, results from similar studies, and other relevant evidence.  Strong results in terms of AUC-ROC and f-beta that reflect cross-validation performance on original development. | 7-10 |
| Implications | 20 | D;V | Discuss the potential clinical use of the model and implications for future research.  Model may have utility in dynamic stratification of patients at risk of mortality in hospital at home setting. | 7-10 |
| Other information | | | | |
| Supplementary information | 21 | D;V | Provide information about the availability of supplementary resources, such as study protocol, Web calculator, and data sets.  Information about baseline patient characteristics, feature analysis, final model hyperparameters, model performance, missingness, SHAP interactions, calibration plots for all the classifiers and TRIPOD guidelines | Supp. material |
| Funding | 22 | D;V | Give the source of funding and the role of the funders for the present study.  This work was Supported by Huma Therapeutics Ltd. The funder had no role in the writing of the manuscript or the decision to submit it for publication. | Title page |

###### **Supplementary Table 5**. TRIPOD Guidelines Report
